# Caffeinated or decaffeinated coffee consumption and risk of cancer incidence: meta-analyses of prospective cohort studies

**DOI:** 10.1101/2023.08.07.23293443

**Authors:** Thanh N. Nguyen, Olga S. Cherepakhin, Devin K. Eng, Masaoki Kawasumi

## Abstract

Many epidemiological studies have investigated the association between coffee consumption and risks for various types of cancer, yet results are conflicting. To determine the impact of coffee consumption on cancer incidence, we systematically reviewed high-quality prospective cohort studies for 10 major cancer types and performed meta-analyses of 63 studies from different countries. For each cancer type, we calculated summary relative risks using the DerSimonian–Laird random-effects model and assessed dose-response relationships. Our meta-analyses found that caffeinated coffee consumption, but not decaffeinated coffee, prevented liver and skin cancers, highlighting the significant role of caffeine in cancer prevention. Furthermore, both caffeinated and decaffeinated coffee prevented endometrial cancer, indicating the role of other active compounds in coffee. Collectively, our meta-analyses revealed that coffee consumption, particularly caffeinated coffee, prevents the incidence of liver, endometrial, and skin cancers in a dose-dependent manner, suggesting that coffee consumption has a large impact on public health.

## Introduction

Cancer is the second leading cause of death in the United States^1^. In 2023, 1,958,310 new cancer cases and 609,820 cancer deaths are projected to occur in the United States^1^. Thus, cancer has a significant impact on public health, and sustainable cancer prevention strategies are needed. Some strategies include avoiding risk factors such as smoking and increasing the consumption of fruits and vegetables^2^. Therefore, identifying which foods and drinks prevent cancer is of great importance.

Coffee is a commonly consumed beverage. In the United States, 49% of adults drink coffee every day^3^. Strikingly, coffee consumption reduces the risk of developing some diseases, including cancer, type 2 diabetes mellitus, and hepatic fibrosis^4–6^. In a large prospective study, coffee consumption was inversely associated with total and cause-specific mortality, showing health benefits of coffee consumption^7^. Coffee contains many bioactive compounds that affect the human body, such as caffeine, chlorogenic acid, diterpenes, and trigonelline^8^. Among these, caffeine has been shown to prevent UV-induced skin carcinogenesis in mouse models^9,10^. Human epidemiological studies also showed that daily consumption of caffeinated coffee, but not decaffeinated coffee, was inversely associated with skin cancer (nonmelanoma skin cancer and melanoma)^11–13^. There are more than 100 types of cancer, which behave differently, and the effect of coffee on cancer prevention may differ among these cancer types. Thus, it is important to comprehensively investigate the preventive effects of caffeinated and decaffeinated coffee on different cancer types.

More than 500 studies have been conducted in different countries to investigate the association between coffee consumption and risks for various types of cancer^4^, yet results are conflicting among different cancer types. Caffeinated coffee consumption prevents skin cancer^11–13^, whereas coffee consumption may increase the risk of bladder cancer^4^. Furthermore, the results from independent studies on the same cancer type may be conflicting; one study showed that coffee intake decreased the risk of ovarian cancer^14^, but another study demonstrated that caffeinated coffee intake increased the risk of ovarian cancer^15^. To better understand the impact of coffee on human cancer incidence, high-quality human studies are needed. Among multiple types of observational studies without any interventions, prospective cohort studies provide the strongest scientific evidence by identifying exposure before the outcome, collecting specific exposure data, and assessing causality^16^. Here, we systematically reviewed prospective cohort studies on major cancer types and performed meta-analyses of 63 studies to quantitatively synthesize the results of multiple independent studies. This allows us to more precisely determine the effects of coffee consumption on cancer incidence than the results derived from individual studies. To elucidate the impact of caffeine, we analyzed the effects of caffeinated and decaffeinated coffee separately. We also assessed whether the effect of coffee on cancer incidence differs between men and women and whether it is dose dependent. Given the fact that coffee is widely consumed and cancer is a common disease, the effect of coffee consumption on cancer incidence could have a large impact on public health.

## Results

### Study selection and characteristics

Through our initial literature search using PubMed, Scopus, and Embase, we found 980 potential articles on coffee consumption and cancer risk (**Supplementary Fig. 1a**). Additionally, 113 articles were found by reviewing reference lists from the initially selected articles and using the “Similar articles” function in PubMed. After uploading these articles to Rayyan QCRI system, duplicates were removed with 861 articles remaining. The initial screening selected only full-text articles, which were then assessed for inclusion criteria. Subsequently, the quality of 74 remaining articles was assessed using the Newcastle-Ottawa Scale (NOS) for nonrandomized studies in meta-analyses, removing seven studies whose NOS scores were lower than 5. After data extraction, we further eliminated four studies from our analysis because the data reported in these four studies were not compatible with other studies for meta-analyses (i.e., the lowest coffee consumption category was not used as reference; the number of cases in the lowest coffee consumption category was less than 10; cancer subtypes were analyzed separately). As a result, 63 studies for 10 major cancer types were included in our meta-analyses.

The 63 prospective cohort studies included 10,068,955 people in total and were conducted in different geographical regions of the world: one study from Australia, 14 from Asia (Japan and Singapore), 21 from Europe (Netherlands, Sweden, Denmark, France, Germany, Greece, Italy, Norway, Spain, United Kingdom, and Finland), and 27 from North America (United States and Canada). The Egger’s test (*p* = 0.0903), Begg’s test (*p* = 0.5424), and a funnel plot of the meta-analysis showed no significant publication bias (**Supplementary Fig. 1b**). The overall summary relative risk (SRR) for 10 major cancer types for high coffee consumption was 0.93 (95% confidence interval [CI], 0.88–0.97) (**Supplementary Fig. 2**). There was high between-study heterogeneity among included cohort studies (*I*^2^ = 65.7%, *p* < 0.001), which prompted us to analyze each cancer type separately.

### Liver cancer (hepatocellular carcinoma)

Overall, our meta-analysis of seven prospective studies^14,17–22^ found that coffee consumption decreased the incidence of hepatocellular carcinoma by 38% (SRR, 0.62; 95% CI, 0.52–0.75) (**Fig. 1a**). Caffeinated coffee consumption reduced the incidence of hepatocellular carcinoma by 46% (SRR, 0.54; 95% CI, 0.39–0.74), whereas decaffeinated coffee consumption had a 16% reduction (SRR, 0.84; 95%CI, 0.64–1.10) (**Fig. 1a**). A greater reduction in the incidence of hepatocellular carcinoma was observed in male coffee drinkers than in female coffee drinkers: 44% reduction in men (SRR, 0.56; 95% CI, 0.39–0.80) and 36% reduction in women (SRR, 0.64; 95% CI, 0.36–1.14) (**Fig. 1b**). A dose-response analysis showed a noticeable decreasing trend in hepatocellular carcinoma incidence with higher coffee consumption: 9.9% reduction per daily cup (**Fig. 1c**).

**Fig. 1:**
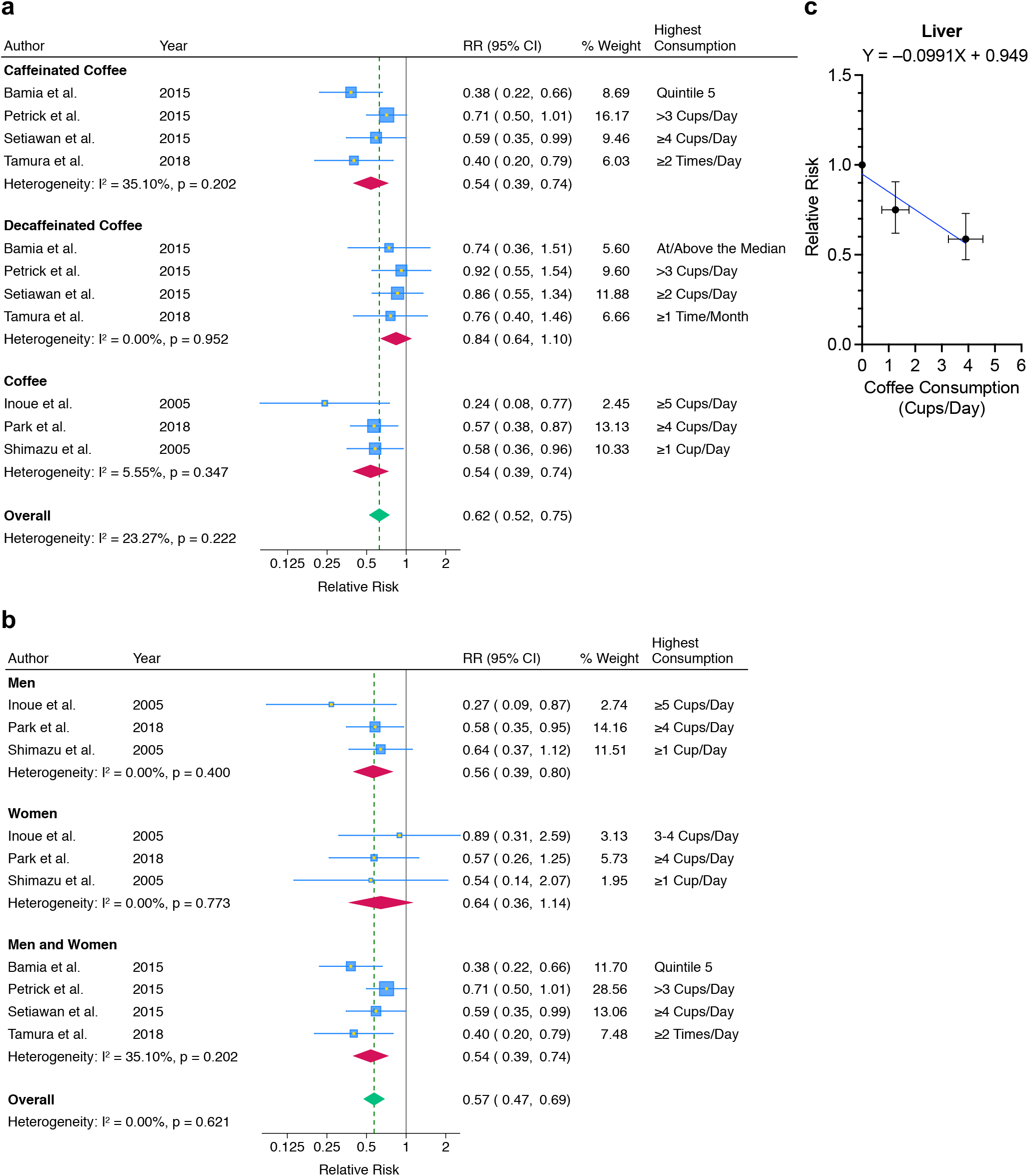
Coffee consumption and hepatocellular carcinoma risk. **a** Meta-analyses classified by coffee type. “Coffee” data did not distinguish between caffeinated and decaffeinated coffee consumption. **b** Meta-analyses classified by sex. Coffee data (without distinguishing between caffeinated and decaffeinated) and caffeinated coffee data, but not decaffeinated coffee data, were included. **c** Dose-response analysis of coffee consumption and hepatocellular carcinoma risk. Mean coffee consumption (cups per day) with 95% confidence interval and summary relative risk with 95% confidence interval for reference, low, and high consumption groups are shown. Coffee data (without distinguishing between caffeinated and decaffeinated) and caffeinated coffee data, but not decaffeinated coffee data, were included.

### Endometrial cancer

Overall, our meta-analysis of 12 prospective studies^14,23–33^ found that coffee consumption decreased endometrial cancer incidence by 30% (SRR, 0.70; 95% CI, 0.61–0.80) (**Fig. 2a**). Endometrial cancer incidence was reduced more in caffeinated coffee drinkers (SRR, 0.61; 95% CI, 0.44–0.84) than in decaffeinated coffee drinkers (SRR, 0.73; 95% CI, 0.58–0.93) (**Fig. 2a**). A dose-response analysis demonstrated a decreasing trend in endometrial cancer incidence with higher coffee consumption: 7.4% reduction per daily cup (**Fig. 2b**).

**Fig. 2:**
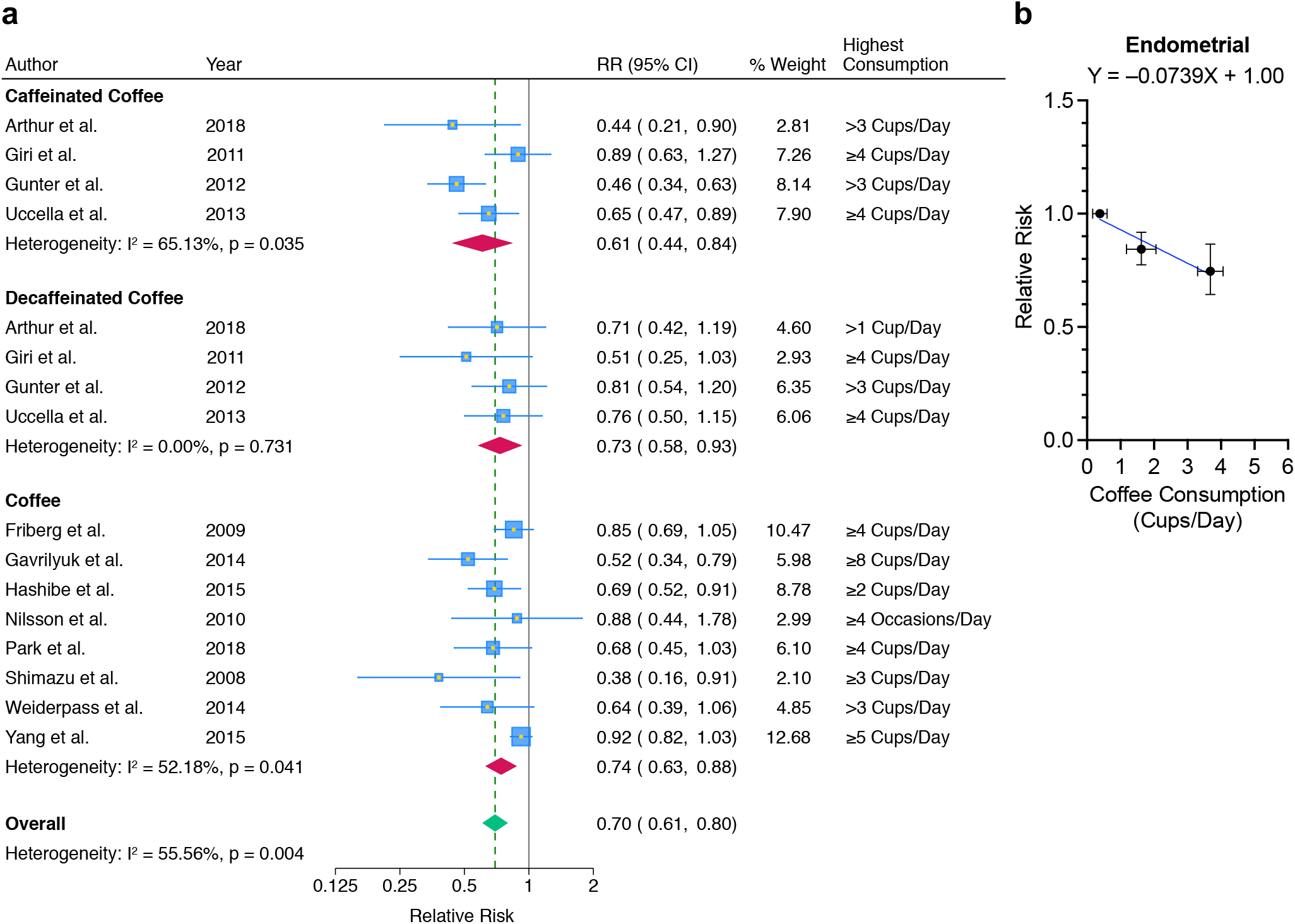
Coffee consumption and endometrial cancer risk. **a** Meta-analyses classified by coffee type. “Coffee” data did not distinguish between caffeinated and decaffeinated coffee consumption. **b** Dose-response analysis of coffee consumption and endometrial cancer risk. Mean coffee consumption (cups per day) with 95% confidence interval and summary relative risk with 95% confidence interval for reference, low, and high consumption groups are shown. Coffee data (without distinguishing between caffeinated and decaffeinated) and caffeinated coffee data, but not decaffeinated coffee data, were included.

### Skin cancer

Overall, our meta-analysis of seven prospective studies^13,14,30,34–37^ found that coffee consumption decreased the incidence of skin cancer by 14% (SRR, 0.86; 95% CI, 0.77–0.96) (**Fig. 3a**). Caffeinated coffee consumption lowered skin cancer incidence by 18% (SRR, 0.82; 95% CI, 0.71– 0.94), whereas decaffeinated coffee consumption had no association with skin cancer incidence (SRR, 0.97; 95% CI, 0.86–1.10) (**Fig. 3a**). There was a greater decrease in skin cancer incidence by 49% in men (SRR, 0.51; 95% CI, 0.20–1.29), compared to a 22% reduction in women (SRR, 0.78; 95% CI, 0.50–1.22) (**Fig. 3b**). A dose-response analysis showed a noticeable decreasing trend in skin cancer incidence with an increase in coffee consumption: 7.8% reduction per daily cup (**Fig. 3c**).

**Fig. 3:**
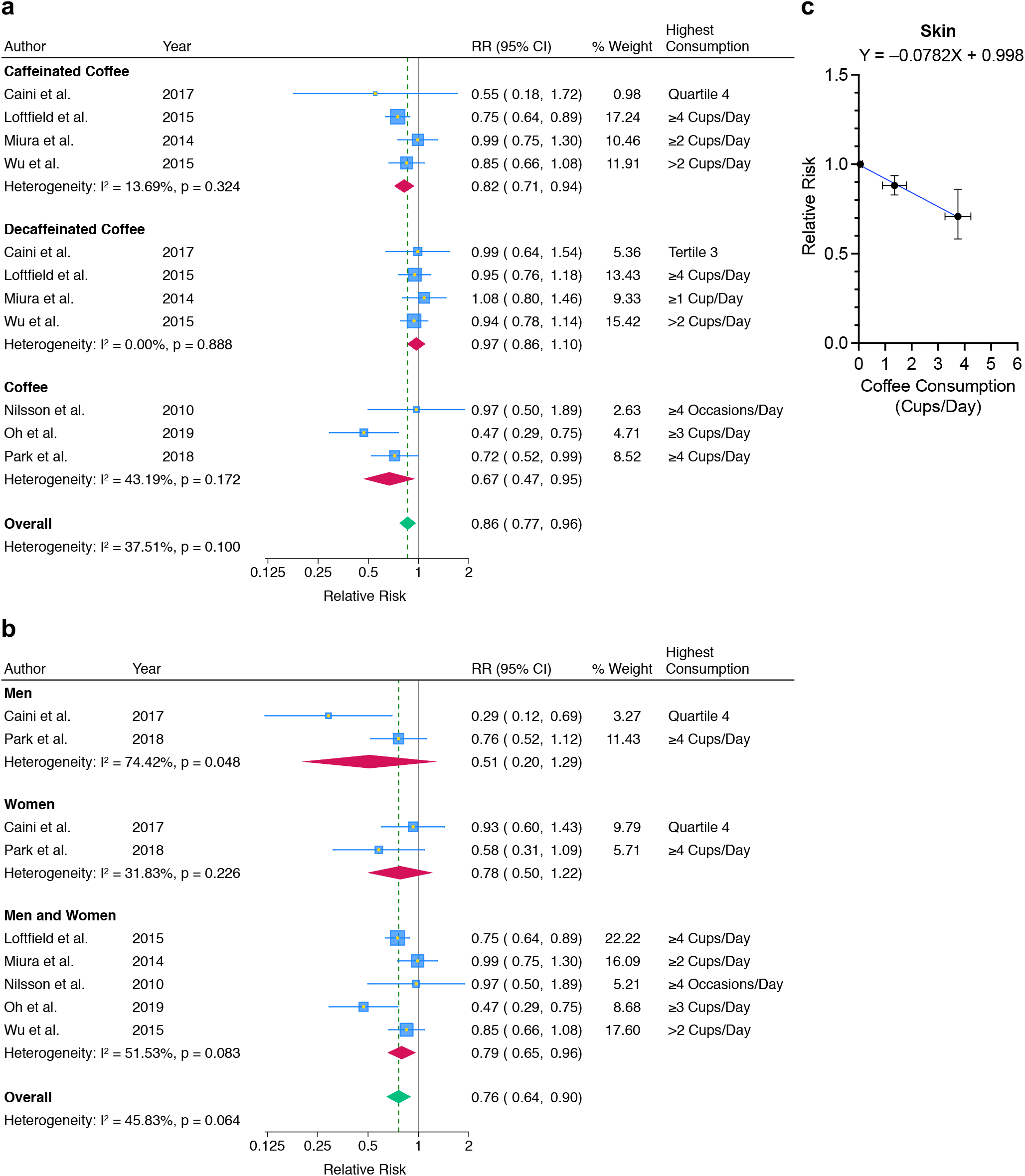
Coffee consumption and skin cancer risk. **a** Meta-analyses classified by coffee type. “Coffee” data did not distinguish between caffeinated and decaffeinated coffee consumption. **b** Meta-analyses classified by sex. Coffee data (without distinguishing between caffeinated and decaffeinated) and caffeinated coffee data, but not decaffeinated coffee data, were included. **c** Dose-response analysis of coffee consumption and skin cancer risk. Mean coffee consumption (cups per day) with 95% confidence interval and summary relative risk with 95% confidence interval for reference, low, and high consumption groups are shown. Coffee data (without distinguishing between caffeinated and decaffeinated) and caffeinated coffee data, but not decaffeinated coffee data, were included.

### Breast cancer

Overall, our meta-analysis of 15 prospective studies^14,23,29,30,38–48^ found that coffee consumption slightly lowered breast cancer incidence (SRR, 0.95; 95% CI, 0.92–0.99) (**Fig. 4a**). Both caffeinated coffee consumption (SRR, 0.95; 95% CI, 0.90–1.01) and decaffeinated coffee consumption (SRR, 0.96; 95% CI, 0.89–1.04) had a decreasing trend in breast cancer risk (**Fig. 4a**). A dose-response analysis showed a decreasing trend in breast cancer incidence with an increase in coffee consumption: 1.2% reduction per daily cup (**Fig. 4b**).

**Fig. 4:**
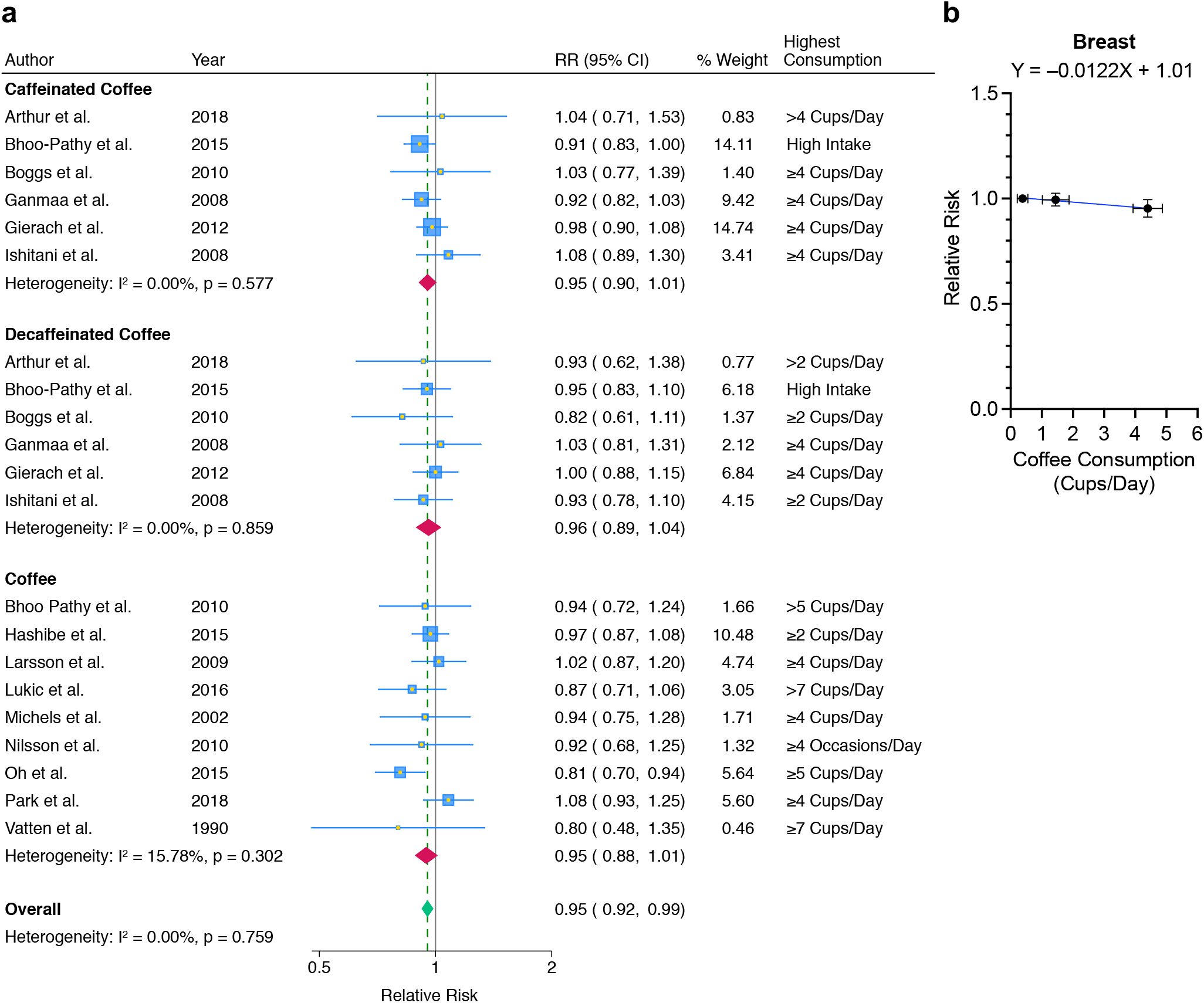
Coffee consumption and breast cancer risk. **a** Meta-analyses classified by coffee type. “Coffee” data did not distinguish between caffeinated and decaffeinated coffee consumption. **b** Dose-response analysis of coffee consumption and breast cancer risk. Mean coffee consumption (cups per day) with 95% confidence interval and summary relative risk with 95% confidence interval for reference, low, and high consumption groups are shown. Coffee data (without distinguishing between caffeinated and decaffeinated) and caffeinated coffee data, but not decaffeinated coffee data, were included.

### Pancreatic cancer

Overall, our meta-analysis of eight prospective studies^14,30,49–54^ found that coffee consumption had no significant association with pancreatic cancer incidence (SRR, 0.96; 95% CI, 0.84–1.09) (**Fig. 5a**). Caffeinated coffee consumption may protect against pancreatic cancer (SRR, 0.94; 95% CI, 0.59–1.48), whereas decaffeinated coffee consumption may increase pancreatic cancer risk (SRR, 1.08; 95% CI, 0.77–1.50) (**Fig. 5a**). Coffee consumption may have a greater protective effect in men (SRR, 0.83; 95% CI 0.62–1.12) than in women (SRR, 0.93; 95% CI 0.76–1.12) (**Fig. 5b**). A dose-response analysis showed a decreasing trend in pancreatic cancer incidence with an increase in coffee consumption (**Fig. 5c**).

**Fig. 5:**
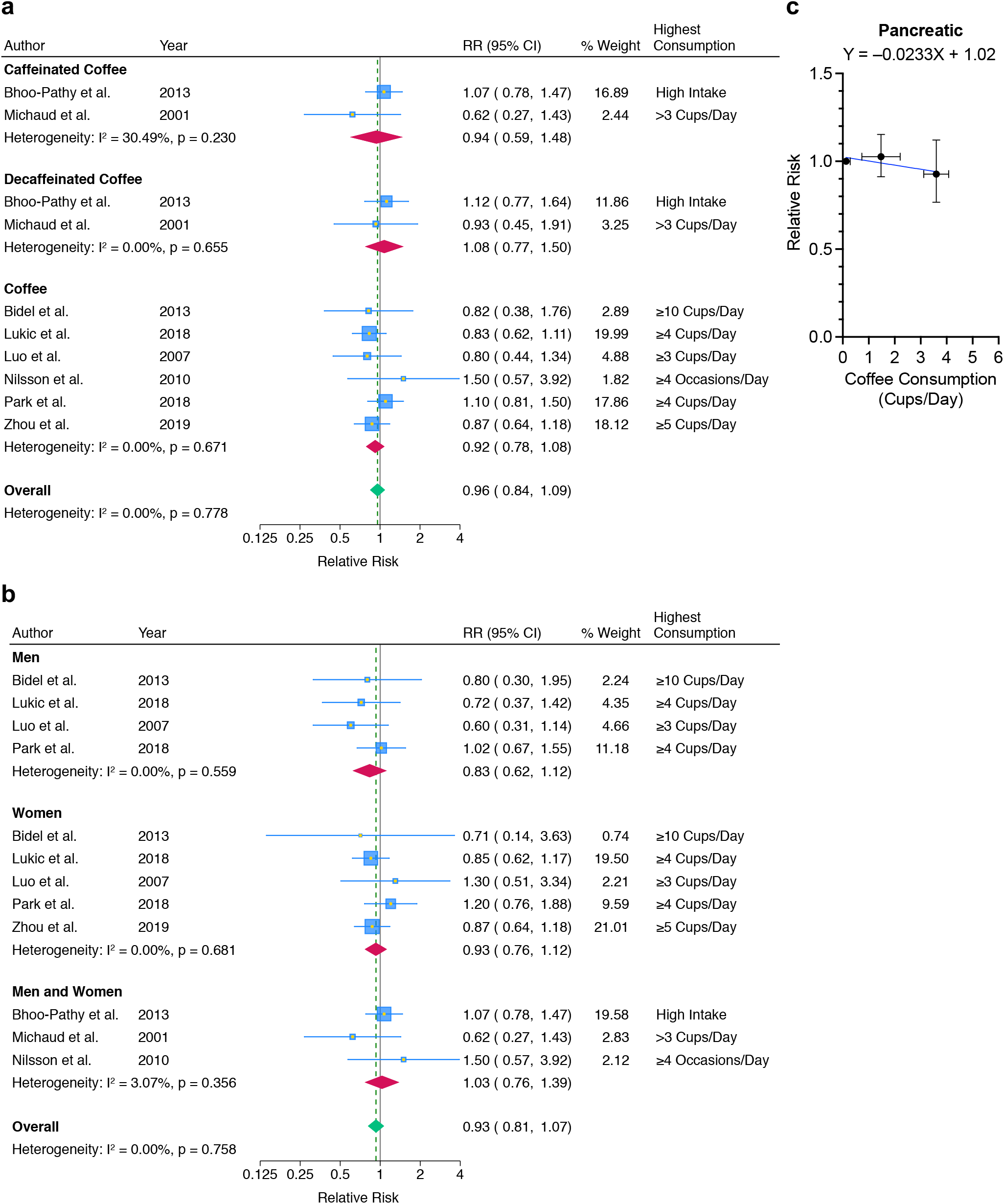
Coffee consumption and pancreatic cancer risk. **a** Meta-analyses classified by coffee type. “Coffee” data did not distinguish between caffeinated and decaffeinated coffee consumption. **b** Meta-analyses classified by sex. Coffee data (without distinguishing between caffeinated and decaffeinated) and caffeinated coffee data, but not decaffeinated coffee data, were included. **c** Dose-response analysis of coffee consumption and pancreatic cancer risk. Mean coffee consumption (cups per day) with 95% confidence interval and summary relative risk with 95% confidence interval for reference, low, and high consumption groups are shown. Coffee data (without distinguishing between caffeinated and decaffeinated) and caffeinated coffee data, but not decaffeinated coffee data, were included.

### Prostate cancer

Our meta-analysis of eight prospective studies^29,30,55–60^ found that the overall SRR for prostate cancer with coffee consumption was 0.98 (95% CI, 0.94–1.02), indicating no significant association between coffee consumption and prostate cancer risk (**Fig. 6a**). Caffeinated (SRR, 0.98; 95% CI, 0.89–1.07) and decaffeinated coffee consumption (SRR, 0.98; 95% CI, 0.87–1.11) also had no association with prostate cancer risk (**Fig. 6a**). A dose-response analysis demonstrated no significant relationship between coffee consumption and prostate cancer risk (**Fig. 6b**).

**Fig. 6:**
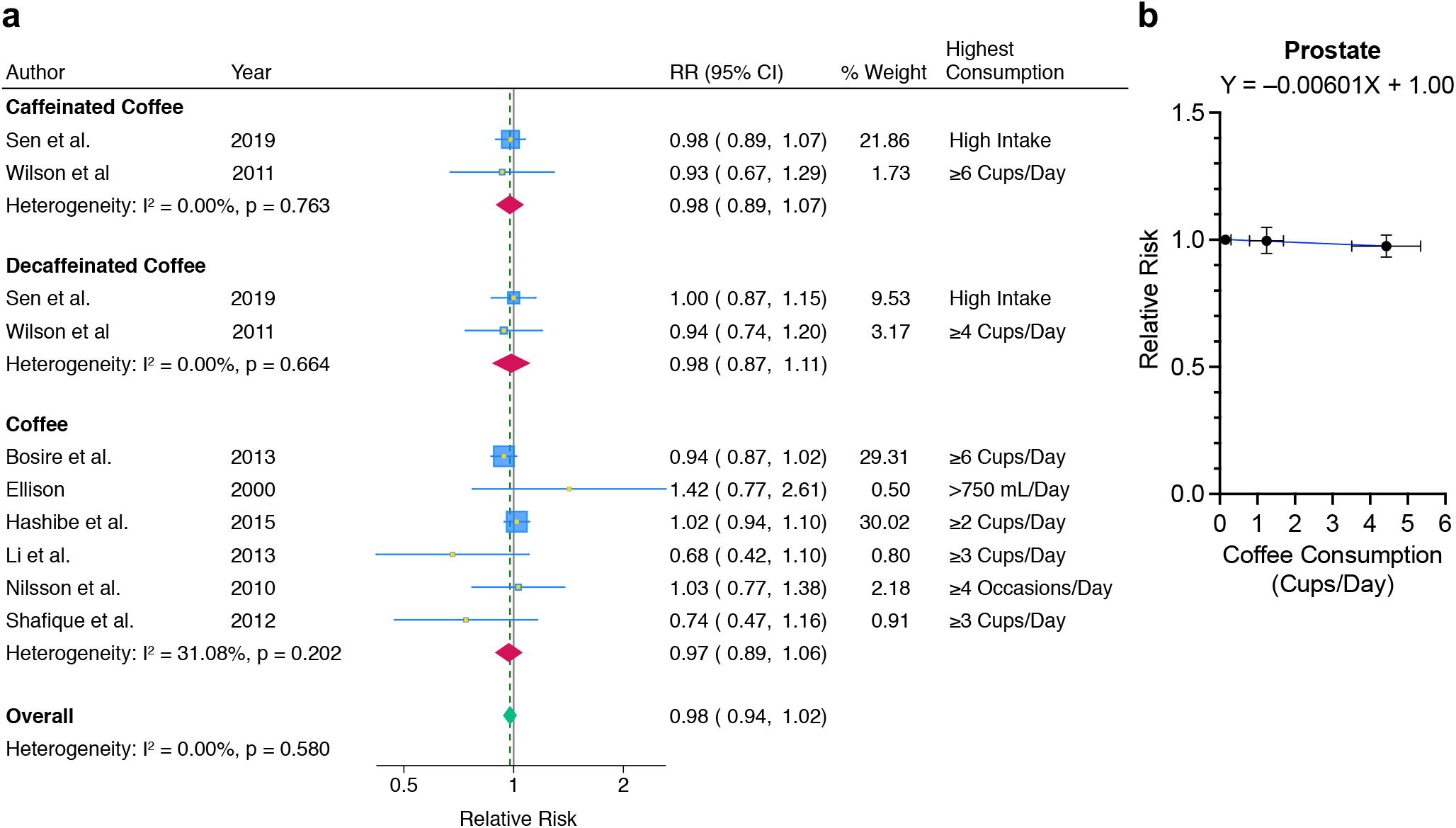
Coffee consumption and prostate cancer risk. **a** Meta-analyses classified by coffee type. “Coffee” data did not distinguish between caffeinated and decaffeinated coffee consumption. **b** Dose-response analysis of coffee consumption and prostate cancer risk. Mean coffee consumption (cups per day) with 95% confidence interval and summary relative risk with 95% confidence interval for reference, low, and high consumption groups are shown. Coffee data (without distinguishing between caffeinated and decaffeinated) and caffeinated coffee data, but not decaffeinated coffee data, were included.

### Colorectal cancer

Overall, our meta-analysis of 11 prospective studies^14,30,45,61–68^ found that coffee consumption had no association with colorectal cancer incidence (SRR, 0.99; 95% CI, 0.91–1.08) (**Fig. 7a**). Intriguingly, decaffeinated coffee consumption had a decreasing trend in colorectal cancer incidence (SRR, 0.88; 95% CI, 0.73–1.07), but caffeinated coffee consumption did not (SRR, 0.99; 95% CI, 0.86–1.14) (**Fig. 7a**). SRRs for male and female coffee drinkers were 1.10 (95% CI, 0.95–1.28) and 1.02 (95% CI, 0.85–1.21), respectively (**Fig. 7b**). A dose-response analysis showed no significant relationship between coffee consumption and colorectal cancer risk (**Fig. 7c**).

**Fig. 7:**
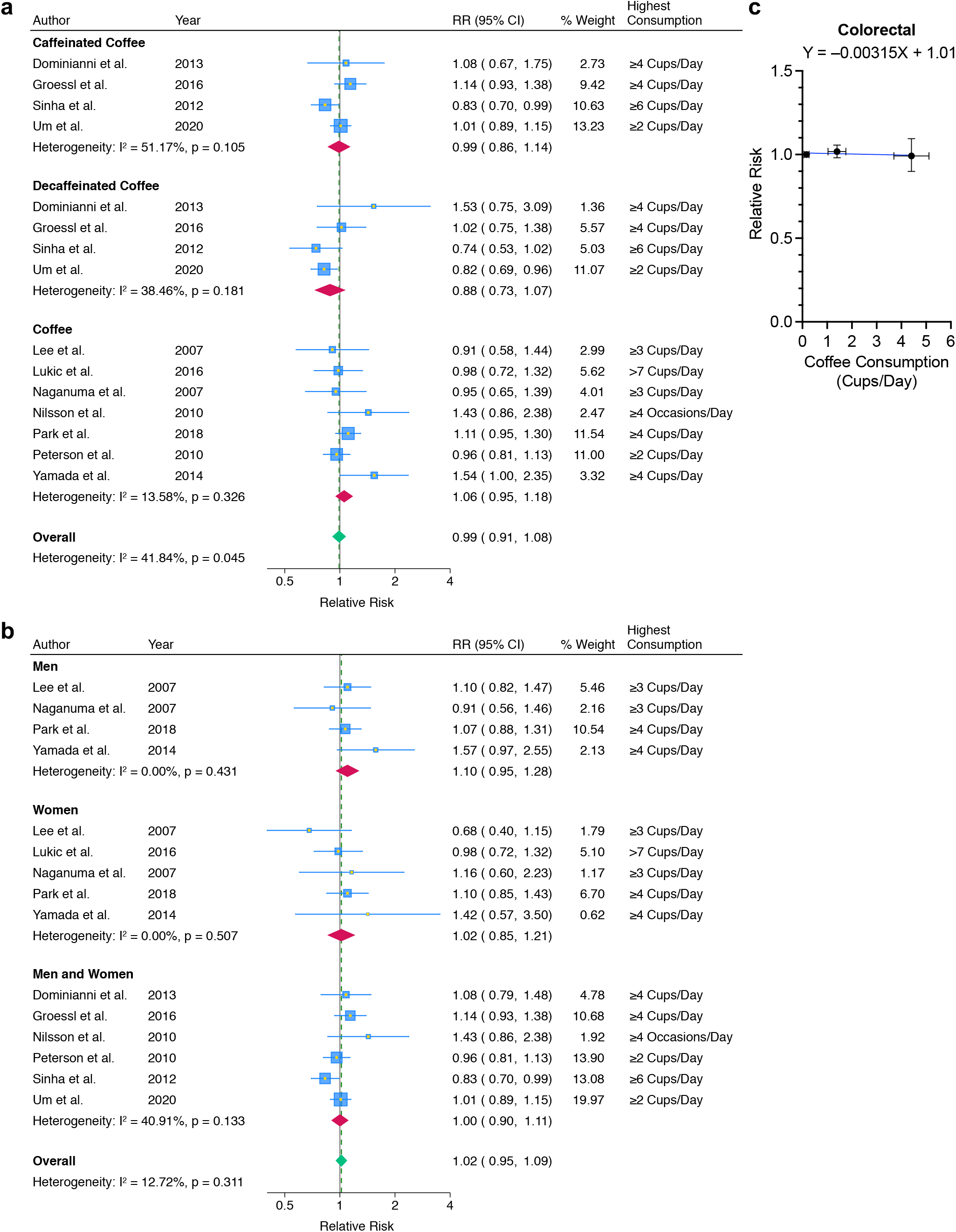
Coffee consumption and colorectal cancer risk. **a** Meta-analyses classified by coffee type. “Coffee” data did not distinguish between caffeinated and decaffeinated coffee consumption. **b** Meta-analyses classified by sex. Coffee data (without distinguishing between caffeinated and decaffeinated) and caffeinated coffee data, but not decaffeinated coffee data, were included. **c** Dose-response analysis of coffee consumption and colorectal cancer risk. Mean coffee consumption (cups per day) with 95% confidence interval and summary relative risk with 95% confidence interval for reference, low, and high consumption groups are shown. Coffee data (without distinguishing between caffeinated and decaffeinated) and caffeinated coffee data, but not decaffeinated coffee data, were included.

### Ovarian cancer

Overall, our meta-analysis of nine prospective studies^14,15,23,29,30,45,69–71^ found that SRR for ovarian cancer in coffee drinkers was 1.00 (95% CI, 0.82–1.23), indicating no association between coffee consumption and ovarian cancer risk (**Fig. 8a**). The *I*^2^ of 53.9% indicated moderate heterogeneity; the Park et al. study showed hazard ratio (HR) of 0.33 (95% CI, 0.17–0.65)^14^, whereas the Silvera et al. study showed HR of 1.62 (95% CI, 0.95–2.75)^71^. Although it was not statistically significant, caffeinated coffee consumption had an increasing trend in ovarian cancer risk (SRR, 1.13; 95% CI, 0.78–1.64), and decaffeinated coffee consumption had a decreasing trend (SRR, 0.89; 95% CI, 0.68–1.17) (**Fig. 8a**). A dose-response analysis demonstrated no significant relationship between coffee consumption and ovarian cancer risk (**Fig. 8b**).

**Fig. 8:**
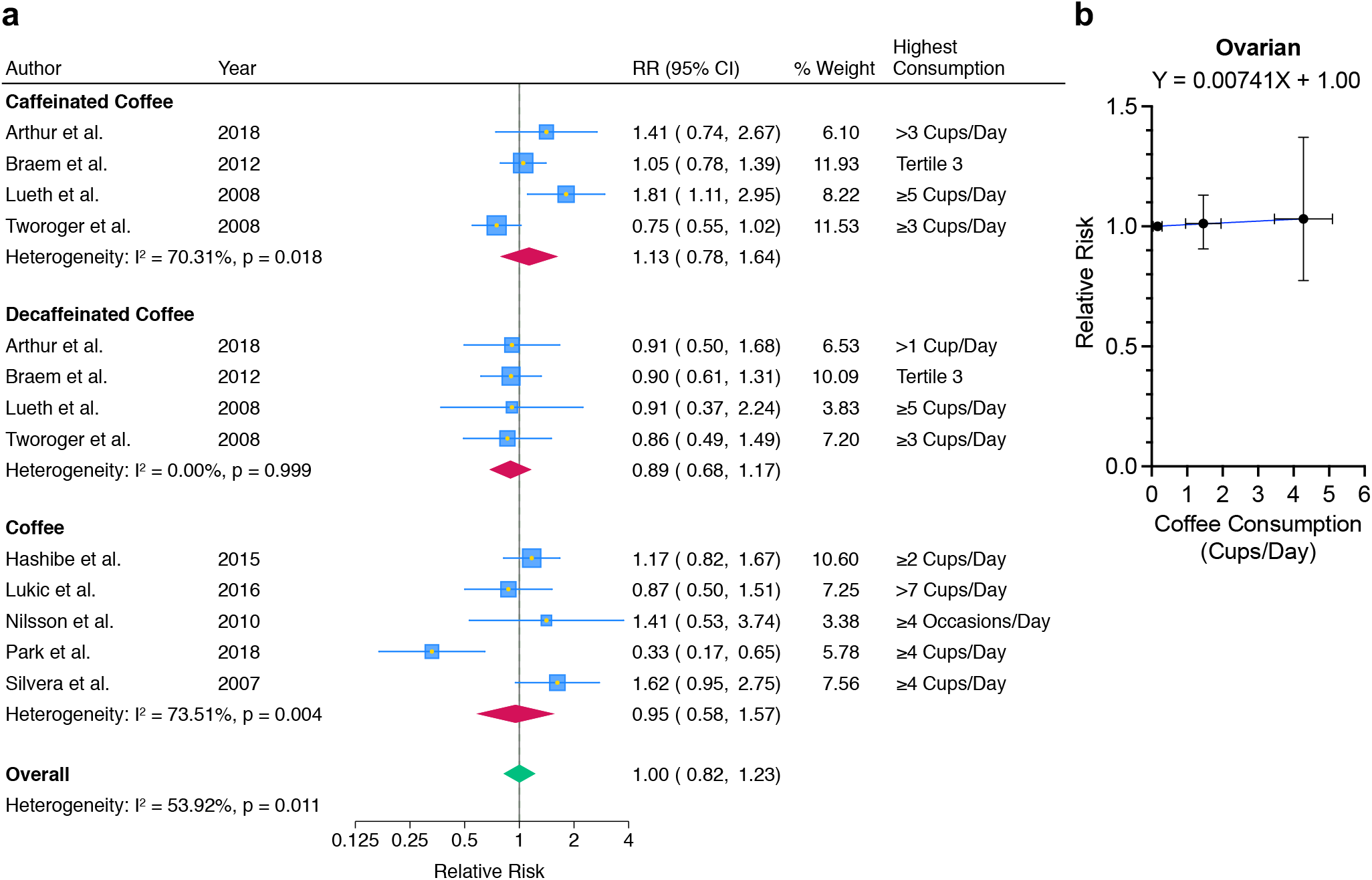
Coffee consumption and ovarian cancer risk. **a** Meta-analyses classified by coffee type. “Coffee” data did not distinguish between caffeinated and decaffeinated coffee consumption. **b** Dose-response analysis of coffee consumption and ovarian cancer risk. Mean coffee consumption (cups per day) with 95% confidence interval and summary relative risk with 95% confidence interval for reference, low, and high consumption groups are shown. Coffee data (without distinguishing between caffeinated and decaffeinated) and caffeinated coffee data, but not decaffeinated coffee data, were included.

### Bladder cancer

Our meta-analysis of six prospective studies^14,29,52,72–74^, including 8,012 bladder cancer cases across the United States, Japan, Norway, and Sweden, found that SRR for bladder cancer in coffee drinkers was 1.11 (95% CI, 0.98–1.26), demonstrating no significant association between coffee consumption and bladder cancer risk (**Fig. 9a**). Only the Loftfield et al. study differentiated between caffeinated and decaffeinated coffee consumption; HRs for caffeinated coffee and decaffeinated coffee were 1.20 (95% CI, 1.06–1.35) and 1.18 (95% CI, 1.00–1.38), respectively, when adjusted for smoking and other confounders^72^. Positive associations between coffee consumption and bladder cancer risk were found among those who had ever smoked, but not in those who had never smoked, suggesting that residual confounding from imperfect measurement of smoking or unmeasured risk factors contributes to the observed positive associations^72^. SRRs for male and female coffee drinkers were 1.08 (95% CI, 0.84–1.39) and 0.96 (95% CI, 0.71–1.29), respectively (**Fig. 9b**). A dose-response analysis showed no significant relationship between coffee consumption and bladder cancer risk (**Fig. 9c**).

**Fig. 9:**
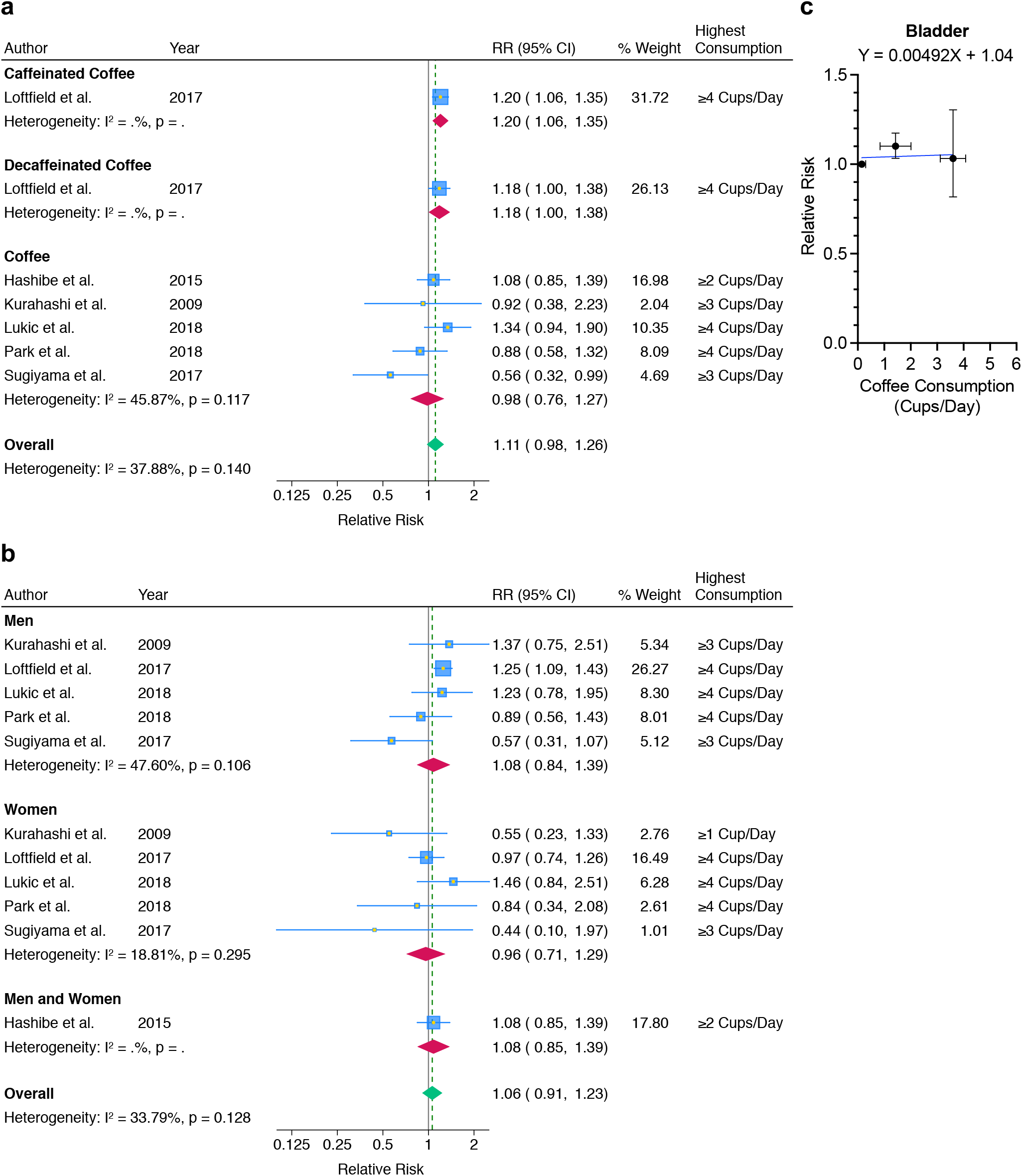
Coffee consumption and bladder cancer risk. **a** Meta-analyses classified by coffee type. “Coffee” data did not distinguish between caffeinated and decaffeinated coffee consumption. **b** Meta-analyses classified by sex. Coffee data (without distinguishing between caffeinated and decaffeinated), but not caffeinated coffee data or decaffeinated coffee data, were included. **c** Dose-response analysis of coffee consumption and bladder cancer risk. Mean coffee consumption (cups per day) with 95% confidence interval and summary relative risk with 95% confidence interval for reference, low, and high consumption groups are shown. Coffee data (without distinguishing between caffeinated and decaffeinated) and caffeinated coffee data, but not decaffeinated coffee data, were included.

### Lung cancer

Our meta-analysis of five prospective studies^14,29,45,75,76^ included 12,725 cases of lung cancer and found an overall SRR of 1.19 (95% CI, 1.07–1.34) for coffee drinkers, demonstrating a significant positive association (**Fig. 10a**). Only the Guertin et al. study differentiated between caffeinated and decaffeinated coffee consumption; HRs for caffeinated coffee and decaffeinated coffee were 1.18 (95% CI, 1.07–1.31) and 1.13 (95% CI, 1.00–1.29), respectively^75^. Compared to SRR for male coffee drinkers (SRR, 1.08; 95% CI, 0.90–1.29), SRR for female coffee drinkers was higher (SRR, 1.48; 95% CI, 0.97–2.28) (**Fig. 10b**). This is due to the Lukic et al. study that found a greater effect for heavy coffee drinkers (>7 cups per day)^45^, which is a higher consumption than in other studies. A dose-response analysis demonstrated an increasing trend in lung cancer incidence with higher coffee consumption: 5.3% increase per daily cup (**Fig. 10c**).

**Fig. 10:**
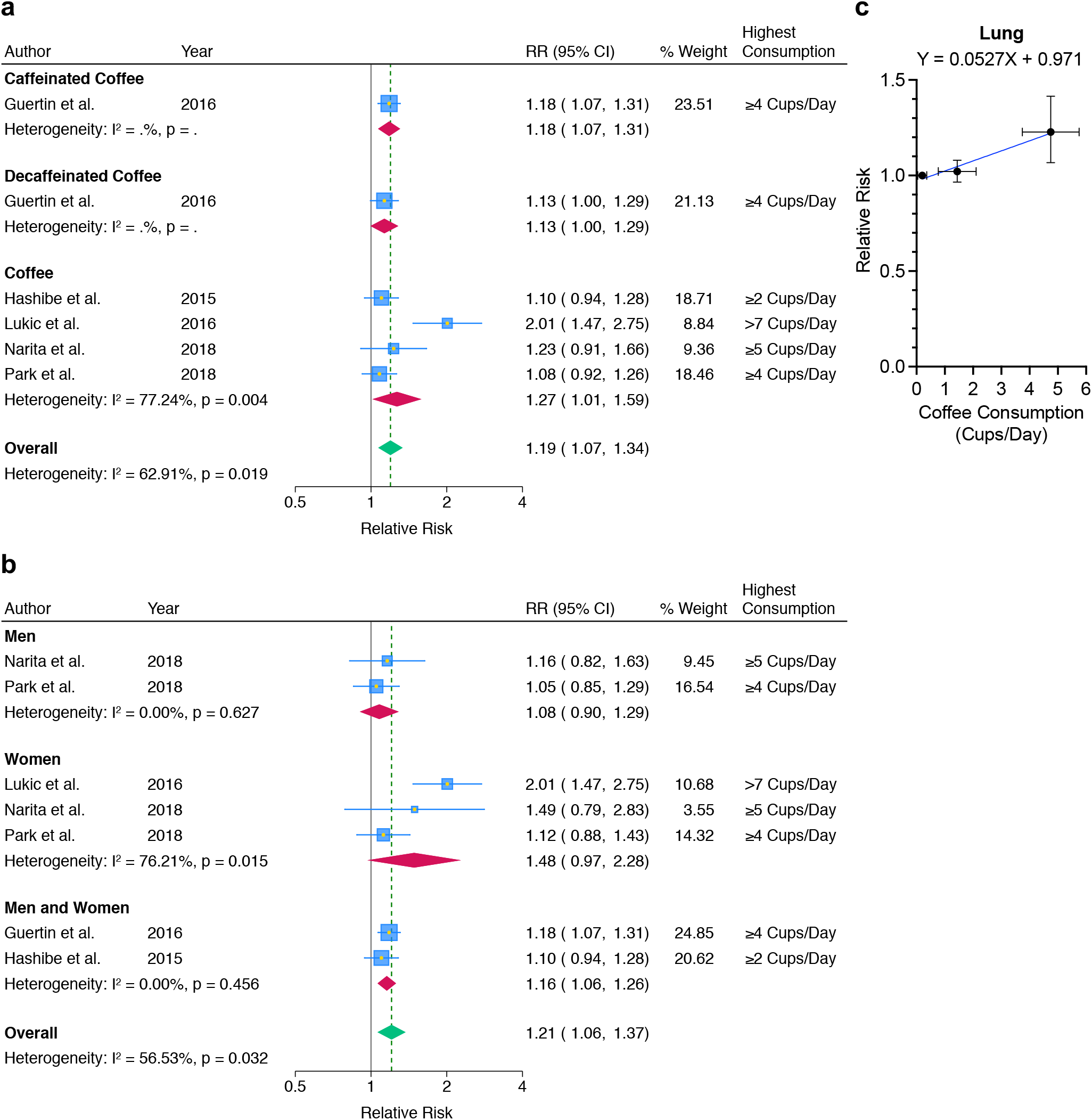
Coffee consumption and lung cancer risk. **a** Meta-analyses classified by coffee type. “Coffee” data did not distinguish between caffeinated and decaffeinated coffee consumption. **b** Meta-analyses classified by sex. Coffee data (without distinguishing between caffeinated and decaffeinated) and caffeinated coffee data, but not decaffeinated coffee data, were included. **c** Dose-response analysis of coffee consumption and lung cancer risk. Mean coffee consumption (cups per day) with 95% confidence interval and summary relative risk with 95% confidence interval for reference, low, and high consumption groups are shown. Coffee data (without distinguishing between caffeinated and decaffeinated) and caffeinated coffee data, but not decaffeinated coffee data, were included.

## Discussion

To investigate whether coffee prevents cancer, we systematically identified and meta-analyzed 63 high-quality prospective cohort studies from different geographical regions of the world, which included 10,068,955 people in total. We excluded case-control studies, which may have recall and selection biases. Unlike most prior meta-analyses, we comprehensively included 10 major cancer types, which cover 59.9% of cancer incidences world-wide^77^, and uniquely differentiated between caffeinated and decaffeinated coffee consumption and between men and women. The overall SRR for 10 major cancer types for high coffee consumption was 0.93 (95% CI, 0.88–0.97), demonstrating the cancer-preventive effects of coffee (**Supplementary Fig. 2**). Between-study heterogeneity among included cohort studies was high (*I*^2^ = 65.7%, *p* < 0.001), and thus we analyzed each cancer type separately. Our meta-analyses revealed that coffee consumption reduced the risk of developing liver cancer (SRR, 0.62; 95% CI, 0.52–0.75), endometrial cancer (SRR, 0.70; 95% CI, 0.61–0.80), skin cancer (SRR, 0.86; 95% CI, 0.77–0.96), and breast cancer (SRR, 0.95; 95% CI, 0.92–0.99), whereas coffee consumption increased lung cancer incidence (SRR, 1.19; 95% CI, 1.07–1.34) (**Supplementary Fig. 3**). There was no significant difference in cancer-preventive effects of coffee between male and female coffee drinkers. The observed positive association for coffee consumption with increased risk of lung cancer is likely due to the residual confounding by tobacco smoking. Coffee drinkers were far more likely to smoke tobacco than nondrinkers^75^. Furthermore, the positive association between coffee consumption and risk of lung cancer was only observed in tobacco smokers, but not in nonsmokers^78,79^.

One of the limitations of our meta-analyses is that, even after differentiating caffeinated and decaffeinated coffee consumption, between-study heterogeneity was significant in caffeinated coffee studies for endometrial cancer (*I*^2^ = 65.1%, *p* = 0.035), colorectal cancer (*I*^2^ = 51.2%, *p* = 0.105), and ovarian cancer (*I*^2^ = 70.3%, *p* = 0.018). This could be due to the inaccuracy of self-reported coffee consumption or changes in coffee intake after study recruitment^80^. Also, considerable heterogeneity exists in coffee cup size and caffeine content per serving^81^. Our meta-analyses included studies from many different countries, and there are different cultural norms for standard size and brewing of coffee. The degree of coffee bean roasting alters the chemical composition of coffee^82^, and coffee brewing methods significantly affect caffeine content^83,84^. Furthermore, the effect of coffee consumption on inflammatory responses is variable in humans; coffee consumption had pro-inflammatory effects in some individuals but anti-inflammatory effects in other individuals^85^.

Our meta-analyses found that caffeinated coffee, but not decaffeinated coffee, prevents liver cancer (SRR, 0.54; 95% CI, 0.39–0.74) and skin cancer (SRR, 0.82; 95% CI, 0.71–0.94), highlighting the significant role of caffeine in cancer prevention (**Supplementary Fig. 3**). Coffee contains many bioactive compounds, but previous mouse studies clearly showed that caffeine is the active compound that prevents UV-induced skin carcinogenesis^9,10^. Caffeine inhibits multiple enzymes, including the ATR kinase that senses UV-induced DNA damage^86^. Genetic inhibition of ATR in skin augments UV-induced apoptosis and suppresses UV-induced skin carcinogenesis, suggesting that ATR inhibition is the relevant mechanism for cancer-preventive effects of caffeine^87^. The augmentation of UV-induced apoptosis by caffeine or ATR inhibition may be beneficial to eliminate DNA-damaged cells that are precancerous, leading to the suppression of UV-induced skin cancer development^88,89^. Additionally, caffeine induces phase II detoxifying and antioxidant enzymes, thereby inhibiting carcinogenesis^90^.

Our meta-analyses also found that decaffeinated coffee prevents endometrial cancer (SRR, 0.73; 95% CI, 0.58–0.93), although caffeinated coffee consumption has a stronger preventive effect (SRR, 0.61; 95% CI, 0.44–0.84) (**Supplementary Fig. 3**). This indicates that active compounds in coffee other than caffeine also play a key role in endometrial cancer prevention. Coffee contains caffeine, chlorogenic acid, diterpenes (mainly cafestol and kahweol), and trigonelline, which affect many different biological processes^8^. Coffee consumption induces glutathione-*S*-transferases (GSTs), which inactivate a broad variety of environmental and dietary toxins^91^. GST induction by unfiltered and paper filtered coffees, differing in cafestol and kahweol contents, was identical, indicating that the diterpene concentrations were not responsible for GST induction^91^. Instead, chlorogenic acid, which is an abundant polyphenol in coffee, stimulates NRF2, a transcription factor that induces antioxidant defense including GST induction^92–94^. Activation of NRF2 decreases reactive oxygen species (ROS) levels and prevents ROS-induced mutations, thereby inhibiting carcinogenesis^88,95^. Cafestol and kahweol have anti-angiogenic properties, which may contribute to cancer prevention^96^.

Due to the popularity of coffee and high frequency of cancer, coffee consumption has a large impact on saving healthcare cost. Based on the estimated numbers of new cancer cases in one year (liver 41,210; uterine corpus 66,200; melanoma 97,610; nonmelanoma skin cancer 5,434,193 [equal incidence rates for basal cell carcinoma and squamous cell carcinoma])^1,97^, per-patient annualized average cancer-attributable costs for initial care (liver $62,776; uterus $39,040; melanoma $8,537; basal cell carcinoma $1,456; squamous cell carcinoma $1,326)^98–100^, and per-cup risk changes from our meta-analyses (liver –9.9%; endometrial –7.4%; skin –7.8%), we estimate that total $1.1 billion of healthcare cost could be saved annually in the United States if Americans drink one additional cup of coffee every day. Furthermore, a previous large prospective cohort study showed that coffee consumption reduced all-cause mortality, highlighting the health benefits of coffee^7^. However, coffee consumption was not inversely associated with deaths due to cancer^7^. This may be because the greatest number of cancer deaths are from cancers of the lung, prostate, colorectum, and pancreas in men and of the lung, breast, colorectum, pancreas, and ovary in women^1^. All these cancers were not prevented by coffee consumption in our meta-analyses, except for a minimal preventive effect found for breast cancer.

Taken together, our meta-analyses of high-quality prospective cohort studies on 10 major cancer types from different countries revealed that coffee consumption, in particular caffeinated coffee, prevents the incidence of liver, endometrial, and skin cancers in a dose-dependent manner. Further investigations are needed to clarify why some types of cancer are prevented by coffee consumption while others are not.

## Methods

### Literature search

We utilized electronic databases PubMed, Scopus, and Embase to comprehensively search for peer-reviewed prospective cohort studies published up to July 2020 without a specific start date. To create a comprehensive list of potential studies, we used the following search terms for all three databases: caffeine, coffee, economics, consumption, drinking, skin cancer, breast cancer, lung cancer, prostate cancer, colorectal cancer, non-Hodgkin lymphoma, renal cell cancer, endometrial cancer, childhood leukemia, pancreatic cancer, thyroid cancer, hepatocellular cancer, ovarian cancer, bladder cancer, epidemiology, prevalence, incidence, risk, humans, and English language. We also utilized reference lists from compiled studies to get a comprehensive list of cohort studies. Furthermore, we utilized the “Similar articles” function in PubMed to include other related articles. Search results were uploaded to Rayyan QCRI system to detect and delete duplications among the three databases.

### Inclusion and exclusion criteria

When compiling a list of potential cohort studies to systematically review, we only included articles that fulfilled the following inclusion criteria: full-length, published, peer-reviewed articles written in English with a prospective cohort study design that included relative risks (or hazard ratios) with 95% confidence intervals (CIs), dose-response relationships between coffee consumption and cancer risk, human subjects, and coffee beverage. Studies were excluded if they are a case-control or retrospective cohort study, do not discuss coffee or caffeine, or do not have a dose-response categorization. In the instance that multiple studies analyzed the same cohort, we included the most updated study.

After a comprehensive list was compiled, we performed a full-text screening to further eliminate any unrelated studies that did not fulfill our inclusion criteria. To assess the quality of prospective cohort studies we selected, we utilized the Newcastle-Ottawa Scale (NOS) for nonrandomized studies in meta-analyses^101^. Studies that had a NOS score less than 5 were excluded. After our comprehensive screening, we had to eliminate our analysis for non-Hodgkin lymphoma, renal cell cancer, childhood leukemia, and thyroid cancer because of limited data availability from prospective cohort studies. As a result, the following 10 cancer types were included in the final analysis: liver, endometrial, skin, breast, pancreatic, prostate, colorectal, ovarian, bladder, and lung.

### Data extraction

From the final list of high-quality prospective cohort studies, we extracted the following data: publication year, cancer type, country, region, study year, total sample size, relative risk (or hazard ratio) with 95% CI for each dose of coffee consumption, coffee type (caffeinated and decaffeinated), and sex (men and women). Only multivariable adjusted data were extracted to eliminate potential confounding factors.

### Data analysis

We performed random-effects meta-analyses using Stata 18 (StataCorp LLC) to integrate the results of included studies for the highest coffee consumption. For each cancer type (liver, endometrial, skin, breast, pancreatic, prostate, colorectal, ovarian, bladder, and lung), coffee type (caffeinated and decaffeinated), and sex (men and women), summary relative risks (SRRs) and 95% CIs were calculated using the DerSimonian–Laird random-effects model, estimating the between-study variance. Hazard ratio is an estimate of relative risk^102^, and thus we treated hazard ratio as relative risk to calculate SRR. To quantify the impact of heterogeneity in a meta-analysis, we calculated *I*^2^, which ranges from 0% to 100%^103^.

For dose-response analyses of coffee consumption and cancer risk, we performed linear regression using Prism 9 (GraphPad Software). Coffee data (without distinguishing between caffeinated and decaffeinated) and caffeinated coffee data, but not decaffeinated coffee data, were included because the majority of coffee drinkers consume caffeinated coffee rather than decaffeinated coffee^104^. Due to the varying nature of how each cohort study reported the amount of coffee consumption, we categorized varying amounts of coffee consumption into low and high consumption groups. We defined low consumption to be less than three cups of coffee per day and high consumption to be greater than or equal to three cups per day. After this grouping, we used relative risk (or hazard ratio) from each study and calculated a SRR for each consumption group. The x-value for our linear regression model represented the mean daily cup of coffee consumption with its 95% CI. The y-value represented SRR with reference set at 1. We excluded the studies that reported nonquantifiable coffee consumption from our linear regression analysis.

Publication bias in our meta-analysis was evaluated using the Egger’s test, Begg’s test, and a funnel plot after applying the trim-and-fill method^105–107^.

## Author contributions

All authors contributed intellectual input and assistance to this study.

T.N.N. and M.K. designed the research.

T.N.N. and D.K.E. conducted literature search and data extraction.

T.N.N. and M.K. analyzed and visualized data.

T.N.N., O.S.C., D.K.E., and M.K. interpreted data and wrote the manuscript.

M.K. supervised the research.

## Data availability

All data are presented within this paper and in the accompanying Supplementary Information.

