## Supplementary Figures 1 to 3 for "Caffeinated or decaffeinated coffee consumption and risk of cancer incidence: meta-analyses of prospective cohort studies"

**This PDF file includes:**

Supplementary Figures 1 to 3

a

b

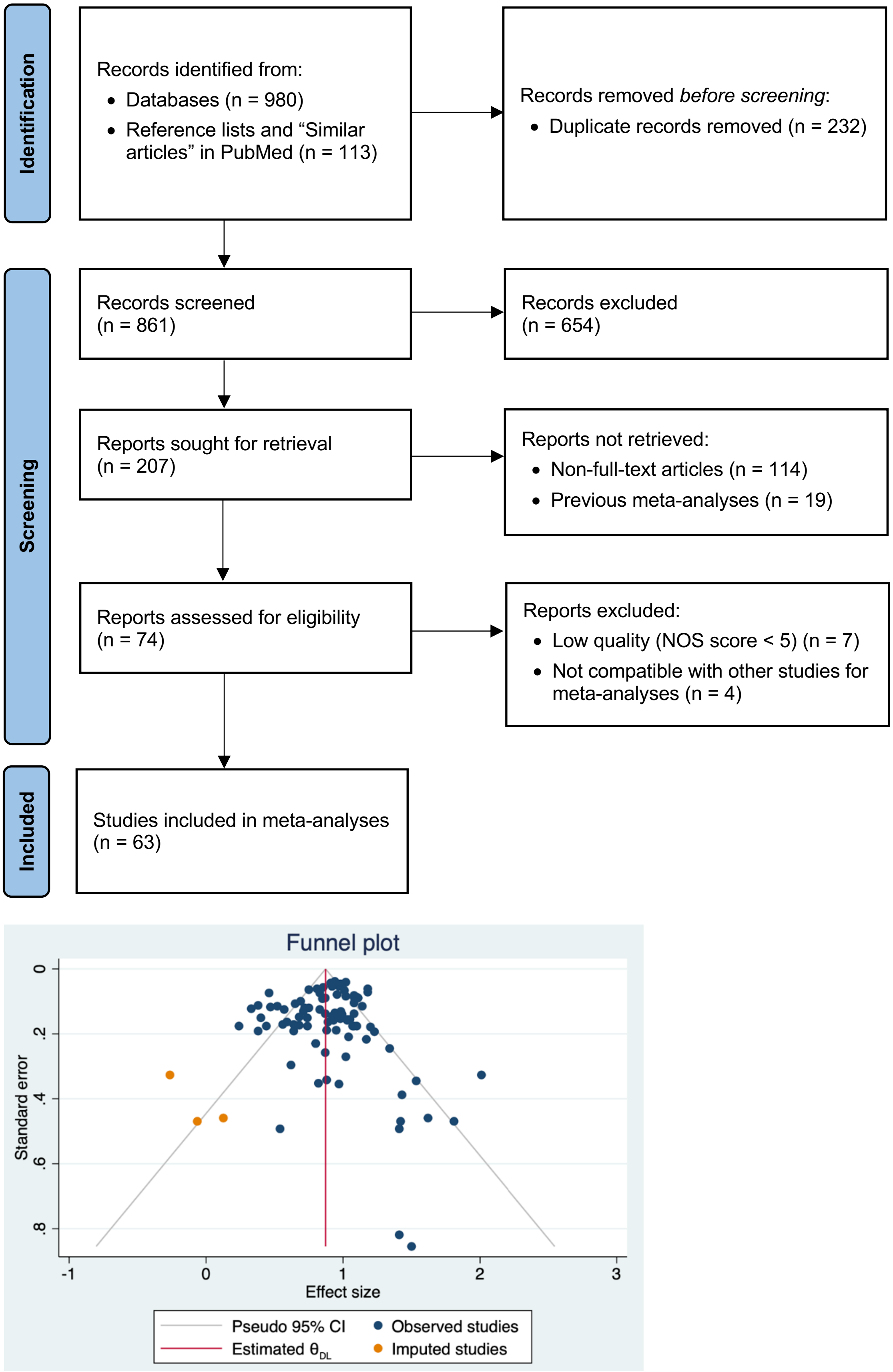


**Supplementary Fig. 1: Systematic review and meta-analysis.**
**a** Systematic review flow diagram.
**b** Funnel plot of the meta-analysis after applying the trim-and-fill method. Each dot represents the standard error (study precision) and effect size of a single study.

| Author | Year | RR (95% CI) | % Weight | Highest Consumption |
|---|---|---|---|---|
| **Liver** | | | | |
| Bamia et al. | 2015 | 0.38 ( 0.22, 0.66) | 0.55 | Quintile 5 |
| Inoue et al. | 2005 | 0.24 ( 0.08, 0.77) | 0.16 | ≥5 Cups/Day |
| Park et al. | 2018 | 0.57 ( 0.38, 0.87) | 0.83 | ≥4 Cups/Day |
| Petrick et al. | 2015 | 0.71 ( 0.50, 1.01) | 1.02 | >3 Cups/Day |
| Setiawan et al. | 2015 | 0.59 ( 0.35, 0.99) | 0.60 | ≥4 Cups/Day |
| Shimazu et al. | 2005 | 0.58 ( 0.36, 0.96) | 0.65 | ≥1 Cup/Day |
| Tamura et al. | 2018 | 0.40 ( 0.20, 0.79) | 0.38 | ≥2 Times/Day |
| Heterogeneity: $I^2$ = 11.66%, p = 0.341 | | 0.55 ( 0.45, 0.67) | | |
| **Endometrial** | | | | |
| Arthur et al. | 2018 | 0.44 ( 0.21, 0.90) | 0.35 | >3 Cups/Day |
| Friberg et al. | 2009 | 0.85 ( 0.69, 1.05) | 1.64 | ≥4 Cups/Day |
| Gavrilyuk et al. | 2014 | 0.52 ( 0.34, 0.79) | 0.81 | ≥8 Cups/Day |
| Giri et al. | 2011 | 0.89 ( 0.63, 1.27) | 1.02 | ≥4 Cups/Day |
| Gunter et al. | 2012 | 0.46 ( 0.34, 0.63) | 1.18 | >3 Cups/Day |
| Hashibe et al. | 2015 | 0.69 ( 0.52, 0.91) | 1.30 | ≥2 Cups/Day |
| Nilsson et al. | 2010 | 0.88 ( 0.44, 1.78) | 0.37 | ≥4 Occasions/Day |
| Park et al. | 2018 | 0.68 ( 0.45, 1.03) | 0.83 | ≥4 Cups/Day |
| Shimazu et al. | 2008 | 0.38 ( 0.16, 0.91) | 0.25 | ≥3 Cups/Day |
| Uccella et al. | 2013 | 0.65 ( 0.47, 0.89) | 1.13 | ≥4 Cups/Day |
| Weiderpass et al. | 2014 | 0.64 ( 0.39, 1.06) | 0.63 | >3 Cups/Day |
| Yang et al. | 2015 | 0.92 ( 0.82, 1.03) | 2.15 | ≥5 Cups/Day |
| Heterogeneity: $I^2$ = 65.80%, p = 0.001 | | 0.68 ( 0.58, 0.81) | | |
| **Skin** | | | | |
| Caini et al. | 2017 | 0.55 ( 0.18, 1.72) | 0.16 | Quartile 4 |
| Loftfield et al. | 2015 | 0.75 ( 0.64, 0.89) | 1.88 | ≥4 Cups/Day |
| Miura et al. | 2014 | 0.99 ( 0.75, 1.30) | 1.32 | ≥2 Cups/Day |
| Nilsson et al. | 2010 | 0.97 ( 0.50, 1.89) | 0.40 | ≥4 Occasions/Day |
| Oh et al. | 2019 | 0.47 ( 0.29, 0.75) | 0.68 | ≥3 Cups/Day |
| Park et al. | 2018 | 0.72 ( 0.52, 0.99) | 1.12 | ≥4 Cups/Day |
| Wu et al. | 2015 | 0.85 ( 0.66, 1.08) | 1.45 | >2 Cups/Day |
| Heterogeneity: $I^2$ = 32.98%, p = 0.176 | | 0.78 ( 0.67, 0.91) | | |
| **Breast** | | | | |
| Arthur et al. | 2018 | 1.04 ( 0.71, 1.53) | 0.91 | >4 Cups/Day |
| Bhoo Pathy et al. | 2010 | 0.94 ( 0.72, 1.24) | 1.33 | >5 Cups/Day |
| Bhoo-Pathy et al. | 2015 | 0.91 ( 0.83, 1.00) | 2.25 | High Intake |
| Boggs et al. | 2010 | 1.03 ( 0.77, 1.39) | 1.23 | ≥4 Cups/Day |
| Ganmaa et al. | 2008 | 0.92 ( 0.82, 1.03) | 2.15 | ≥4 Cups/Day |
| Gierach et al. | 2012 | 0.98 ( 0.90, 1.08) | 2.26 | ≥4 Cups/Day |
| Hashibe et al. | 2015 | 0.97 ( 0.87, 1.08) | 2.18 | ≥2 Cups/Day |
| Ishitani et al. | 2008 | 1.08 ( 0.89, 1.30) | 1.75 | ≥4 Cups/Day |
| Larsson et al. | 2009 | 1.02 ( 0.87, 1.20) | 1.91 | ≥4 Cups/Day |
| Lukic et al. | 2016 | 0.87 ( 0.71, 1.06) | 1.69 | >7 Cups/Day |
| Michels et al. | 2002 | 0.94 ( 0.75, 1.28) | 1.35 | ≥4 Cups/Day |
| Nilsson et al. | 2010 | 0.92 ( 0.68, 1.25) | 1.19 | ≥4 Occasions/Day |
| Oh et al. | 2015 | 0.81 ( 0.70, 0.94) | 1.98 | ≥5 Cups/Day |
| Park et al. | 2018 | 1.08 ( 0.93, 1.25) | 1.98 | ≥4 Cups/Day |
| Vatten et al. | 1990 | 0.80 ( 0.48, 1.35) | 0.61 | ≥7 Cups/Day |
| Heterogeneity: $I^2$ = 0.00%, p = 0.501 | | 0.95 ( 0.91, 0.99) | | |
| **Pancreatic** | | | | |
| Bhoo-Pathy et al. | 2013 | 1.07 ( 0.78, 1.47) | 1.14 | High Intake |
| Bidel et al. | 2013 | 0.82 ( 0.38, 1.76) | 0.32 | ≥10 Cups/Day |
| Lukic et al. | 2018 | 0.83 ( 0.62, 1.11) | 1.25 | ≥4 Cups/Day |
| Luo et al. | 2007 | 0.80 ( 0.44, 1.34) | 0.49 | ≥3 Cups/Day |
| Michaud et al. | 2001 | 0.62 ( 0.27, 1.43) | 0.27 | >3 Cups/Day |
| Nilsson et al. | 2010 | 1.50 ( 0.57, 3.92) | 0.21 | ≥4 Occasions/Day |
| Park et al. | 2018 | 1.10 ( 0.81, 1.50) | 1.18 | ≥4 Cups/Day |
| Zhou et al. | 2019 | 0.87 ( 0.64, 1.18) | 1.19 | ≥5 Cups/Day |
| Heterogeneity: $I^2$ = 0.00%, p = 0.677 | | 0.94 ( 0.81, 1.08) | | |
| **Prostate** | | | | |
| Bosire et al. | 2013 | 0.94 ( 0.87, 1.02) | 2.31 | ≥6 Cups/Day |
| Ellison | 2000 | 1.42 ( 0.77, 2.61) | 0.46 | >750 mL/Day |
| Hashibe et al. | 2015 | 1.02 ( 0.94, 1.10) | 2.31 | ≥2 Cups/Day |
| Li et al. | 2013 | 0.68 ( 0.42, 1.10) | 0.67 | ≥3 Cups/Day |
| Nilsson et al. | 2010 | 1.03 ( 0.77, 1.38) | 1.24 | ≥4 Occasions/Day |
| Sen et al. | 2019 | 0.98 ( 0.89, 1.07) | 2.26 | High Intake |
| Shafique et al. | 2012 | 0.74 ( 0.47, 1.16) | 0.73 | ≥3 Cups/Day |
| Wilson et al | 2011 | 0.93 ( 0.67, 1.29) | 1.10 | ≥6 Cups/Day |
| Heterogeneity: $I^2$ = 4.71%, p = 0.394 | | 0.98 ( 0.93, 1.02) | | |
| **Colorectal** | | | | |
| Dominianni et al. | 2013 | 1.08 ( 0.67, 1.75) | 0.67 | ≥4 Cups/Day |
| Groessl et al. | 2016 | 1.14 ( 0.93, 1.38) | 1.71 | ≥4 Cups/Day |
| Lee et al. | 2007 | 0.91 ( 0.58, 1.44) | 0.73 | ≥3 Cups/Day |
| Lukic et al. | 2016 | 0.98 ( 0.72, 1.32) | 1.20 | >7 Cups/Day |
| Naganuma et al. | 2007 | 0.95 ( 0.65, 1.39) | 0.92 | ≥3 Cups/Day |
| Nilsson et al. | 2010 | 1.43 ( 0.86, 2.38) | 0.62 | ≥4 Occasions/Day |
| Park et al. | 2018 | 1.11 ( 0.95, 1.30) | 1.93 | ≥4 Cups/Day |
| Peterson et al. | 2010 | 0.96 ( 0.81, 1.13) | 1.87 | ≥2 Cups/Day |
| Sinha et al. | 2012 | 0.83 ( 0.70, 0.99) | 1.84 | ≥6 Cups/Day |
| Um et al. | 2020 | 1.01 ( 0.89, 1.15) | 2.08 | ≥2 Cups/Day |
| Yamada et al. | 2014 | 1.54 ( 1.00, 2.35) | 0.79 | ≥4 Cups/Day |
| Heterogeneity: $I^2$ = 28.82%, p = 0.171 | | 1.02 ( 0.94, 1.11) | | |
| **Ovarian** | | | | |
| Arthur et al. | 2018 | 1.41 ( 0.74, 2.67) | 0.43 | >3 Cups/Day |
| Braem et al. | 2012 | 1.05 ( 0.78, 1.39) | 1.26 | Tertile 3 |
| Hashibe et al. | 2015 | 1.17 ( 0.82, 1.67) | 1.00 | ≥2 Cups/Day |
| Lueth et al. | 2008 | 1.81 ( 1.11, 2.95) | 0.66 | ≥5 Cups/Day |
| Lukic et al. | 2016 | 0.87 ( 0.50, 1.51) | 0.54 | >7 Cups/Day |
| Nilsson et al. | 2010 | 1.41 ( 0.53, 3.74) | 0.20 | ≥4 Occasions/Day |
| Park et al. | 2018 | 0.33 ( 0.17, 0.65) | 0.40 | ≥4 Cups/Day |
| Silvera et al. | 2007 | 1.62 ( 0.95, 2.75) | 0.58 | ≥4 Cups/Day |
| Tworoger et al. | 2008 | 0.75 ( 0.55, 1.02) | 1.17 | ≥3 Cups/Day |
| Heterogeneity: $I^2$ = 68.26%, p = 0.001 | | 1.05 ( 0.79, 1.39) | | |
| **Bladder** | | | | |
| Hashibe et al. | 2015 | 1.08 ( 0.85, 1.39) | 1.45 | ≥2 Cups/Day |
| Kurahashi et al. | 2009 | 0.92 ( 0.38, 2.23) | 0.24 | ≥3 Cups/Day |
| Loftfield et al. | 2017 | 1.20 ( 1.06, 1.35) | 2.12 | ≥4 Cups/Day |
| Lukic et al. | 2018 | 1.34 ( 0.94, 1.90) | 1.01 | ≥4 Cups/Day |
| Park et al. | 2018 | 0.88 ( 0.58, 1.32) | 0.83 | ≥4 Cups/Day |
| Sugiyama et al. | 2017 | 0.56 ( 0.32, 0.99) | 0.53 | ≥3 Cups/Day |
| Heterogeneity: $I^2$ = 47.49%, p = 0.090 | | 1.06 ( 0.88, 1.28) | | |
| **Lung** | | | | |
| Guertin et al. | 2016 | 1.18 ( 1.07, 1.31) | 2.22 | ≥4 Cups/Day |
| Hashibe et al. | 2015 | 1.10 ( 0.94, 1.28) | 1.94 | ≥2 Cups/Day |
| Lukic et al. | 2016 | 2.01 ( 1.47, 2.75) | 1.16 | >7 Cups/Day |
| Narita et al. | 2018 | 1.23 ( 0.91, 1.66) | 1.21 | ≥5 Cups/Day |
| Park et al. | 2018 | 1.08 ( 0.92, 1.26) | 1.93 | ≥4 Cups/Day |
| Heterogeneity: $I^2$ = 69.67%, p = 0.010 | | 1.22 ( 1.06, 1.42) | | |
| **Overall** | | 0.93 ( 0.88, 0.97) | | |
| Heterogeneity: $I^2$ = 65.73%, p = 0.000 | | | | |

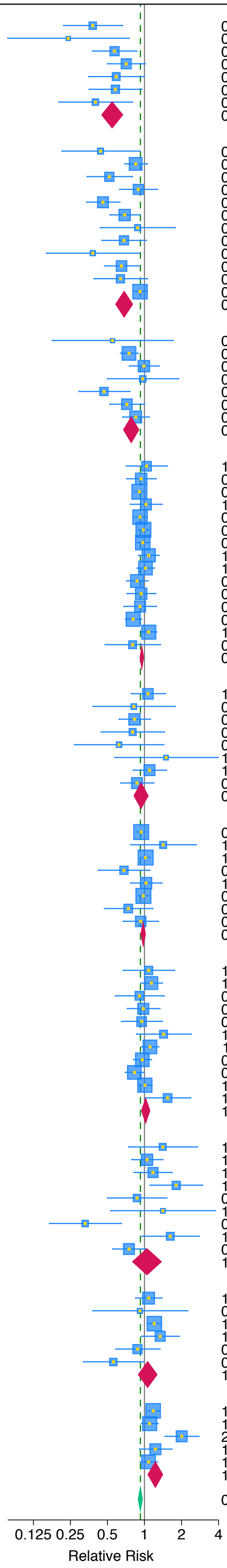


**Supplementary Fig. 2: Summary relative risks of 10 major cancer types for coffee consumption.** Meta-analyses were performed combining coffee data (without distinguishing between caffeinated and decaffeinated) and caffeinated coffee data, but not including decaffeinated coffee data.

| Cancer Type | Summary Relative Risk (95% Confidence Interval) | | | | | Per-Cup Risk Change |
|---|---|---|---|---|---|---|
| | Overall Coffee | Caffeinated | Decaffeinated | Men | Women | |
| Liver | **0.62 * (0.52, 0.75)** | **0.54 * (0.39, 0.74)** | 0.84 (0.64, 1.10) | **0.56 * (0.39, 0.80)** | 0.64 (0.36, 1.14) | **–9.9% [a]** |
| Endometrial | **0.70 * (0.61, 0.80)** | **0.61 * (0.44, 0.84)** | **0.73 * (0.58, 0.93)** | NA | NA | **–7.4% [a]** |
| Skin | **0.86 * (0.77, 0.96)** | **0.82 * (0.71, 0.94)** | 0.97 (0.86, 1.10) | 0.51 (0.20, 1.29) | 0.78 (0.50, 1.22) | **–7.8% [a]** |
| Breast | **0.95 * (0.92, 0.99)** | 0.95 (0.90, 1.01) | 0.96 (0.89, 1.04) | NA | NA | **–1.2% [a]** |
| Pancreatic | 0.96 (0.84, 1.09) | 0.94 (0.59, 1.48) | 1.08 (0.77, 1.50) | 0.83 (0.62, 1.12) | 0.93 (0.76, 1.12) | –2.3% |
| Prostate | 0.98 (0.94, 1.02) | 0.98 (0.89, 1.07) | 0.98 (0.87, 1.11) | NA | NA | –0.6% |
| Colorectal | 0.99 (0.91, 1.08) | 0.99 (0.86, 1.14) | 0.88 (0.73, 1.07) | 1.10 (0.95, 1.28) | 1.02 (0.85, 1.21) | –0.3% |
| Ovarian | 1.00 (0.82, 1.23) | 1.13 (0.78, 1.64) | 0.89 (0.68, 1.17) | NA | NA | +0.7% |
| Bladder | 1.11 (0.98, 1.26) | ND | ND | 1.08 (0.84, 1.39) | 0.96 (0.71, 1.29) | +0.5% |
| Lung | **1.19 * (1.07, 1.34)** | ND | ND | 1.08 (0.90, 1.29) | 1.48 (0.97, 2.28) | **+5.3% [a]** |

**Supplementary Fig. 3: Summary relative risks of 10 major cancer types for coffee consumption and per-cup risk changes.**
"Overall Coffee" includes all coffee data: coffee data (without distinguishing between caffeinated and decaffeinated), caffeinated coffee data, and decaffeinated coffee data. Per-cup risk change is derived from the slope of linear regression in a dose-response analysis. Blue and red highlights indicate statistically significant decrease and increase in cancer risk, respectively. * $p < 0.05$. [a] $p < 0.05$ for high coffee consumption. NA, sex comparison was not available. ND, summary relative risk was not determined because only one study was available.
